# Hormone replacement therapy and elevated breast cancer risk: An artifact of growth acceleration?

**DOI:** 10.1101/2020.04.04.20050708

**Authors:** Jutta Engel, Gabriele Schubert-Fritschle, Dieter Hölzel

## Abstract

**Background:** Available data on accelerated proliferation and increased breast cancer risk due to hormone replacement therapy (HT) are inconsistent. Data on long-term effects of HT are limited. The interaction between several key factors was examined using a model-based approach.

**Methods:** Cohorts of 50 year old women, BCs were randomly generated for 30 years based on the age-specific incidence. A control group received a HT that increased the growth of occult BCs. In a 3rd cohort BCs were additionally induced by HT. This model illustrates the interrelationship of important parameters and allows the simulation and comparison of previously published clinical studies.

**Results:** Using plausible parameters for BC growth factor (GF) and HT-related effects it was demonstrated that HT caused accelerated growth of occult BCs with an apparent increase in incidence and shortened time to diagnosis. The Women’s Health Initiative (WHI) study was reconstructed assuming a GF of 1.43 induced by HT. The decision of millions of women to discontinue or forego HT based on the published risks of the WHI-study in 2002 could explain the marked jump of 6.7% in incidence within a few months. If additional BCs were induced by HT, then these BCs may become apparent after 10 or more years together with those appearing according to the normal incidence. At this time conclusive data on type, timing, and molecular characteristics of HT induced BCs are not yet available.

**Conclusion:** The acceleration in growth of occult BC has been underestimated. Initially HTs can cause an apparent increase in BC incidence thereby explaining the WHI-dependent decrease in 2003. A HT associated BC risk should only be detectable with a delay of ten and more years.

## Introduction

Hormone replacement therapy, hereafter referred to as hormone therapy (HT), has been the subject of over 50 years of research and discussions leading to a varying use in clinical practice ^1^. The reasons for this disparity are reported negative and positive side effects of HTs^2^ such as a higher risk for invasive breast cancer (BC), endometrial cancer, or stroke. On the other hand positive side effects have also been published such as a reduced risk for fractures, diabetes, colon cancer, or heart disease. Cost-benefit analyses are complex due to the dependency on age, the type and duration of HT application. Guidelines generally emphasize the need and the benefit of HTs, but also vary in their restrictiveness. A recently updated meta-analysis ^3,4^ continues this HT narrative by which approximately 5% or 1 million BCs have been induced by HT since 1990. This result of the study motivated the analysis herein. The main objective was to assess the association of BC with key factors using models based on studies and populations, compare their results, and discuss the use and suitability of the currently available data.

## Methods

The models described below were based on cohorts of 50 years old women. BCs were randomly generated for 30 years according to the age-specific incidence^5,6^. For a comparison with published clinical trials the respective age distributions and competing mortality according to life expectancy were considered. The second cohort begins with HT application, and expected effects of HTs were modeled according to 4 parameters: duration of BC growth (GD), duration of HT application (HTD), and an accelerated growth (GF) due to HT. In the 3rd cohort a BC risk was also modeled. It was assumed that proliferation and BC risk begins and ends with HT application, as the well-known BC incidence jump in 2003 suggests.

The modeling approach based on the association of 15 years of GD, 4 years HTD, a doubled GF, and a BC risk is clarified in figure 1. Herein illustrated are the growth trajectories for BCs, the number of tumor cells (log scale), as well as the age of the occult BCs over time. The logarithm of the tumor cells resulted in a linear trajectory for the assumed exponential BC growth. A second key component was the GF, a linear transformation of the growth induced by HT. The impact of growth acceleration may be subdivided into 6 phases (Figure 1B). Plausible model assumptions were taken from peer-reviewed literature, direct comparisons were taken from the Women’s Health Initiative Study (WHI- S)^7,8^, and the updated meta-analysis.^3^ The model was constructed and all statistical analysis were done in R Version 3.1.3.^9^

**Fig. 1:**
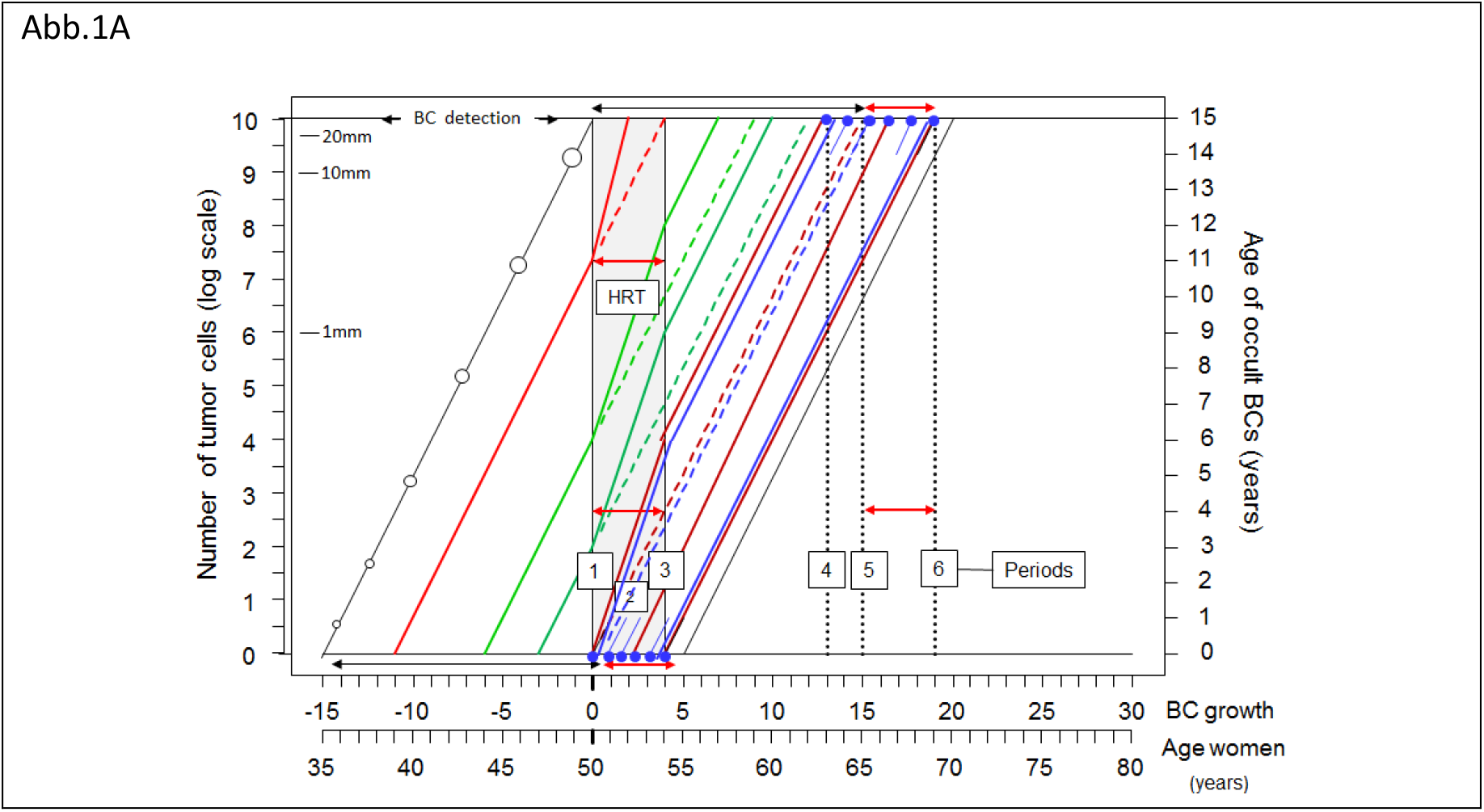
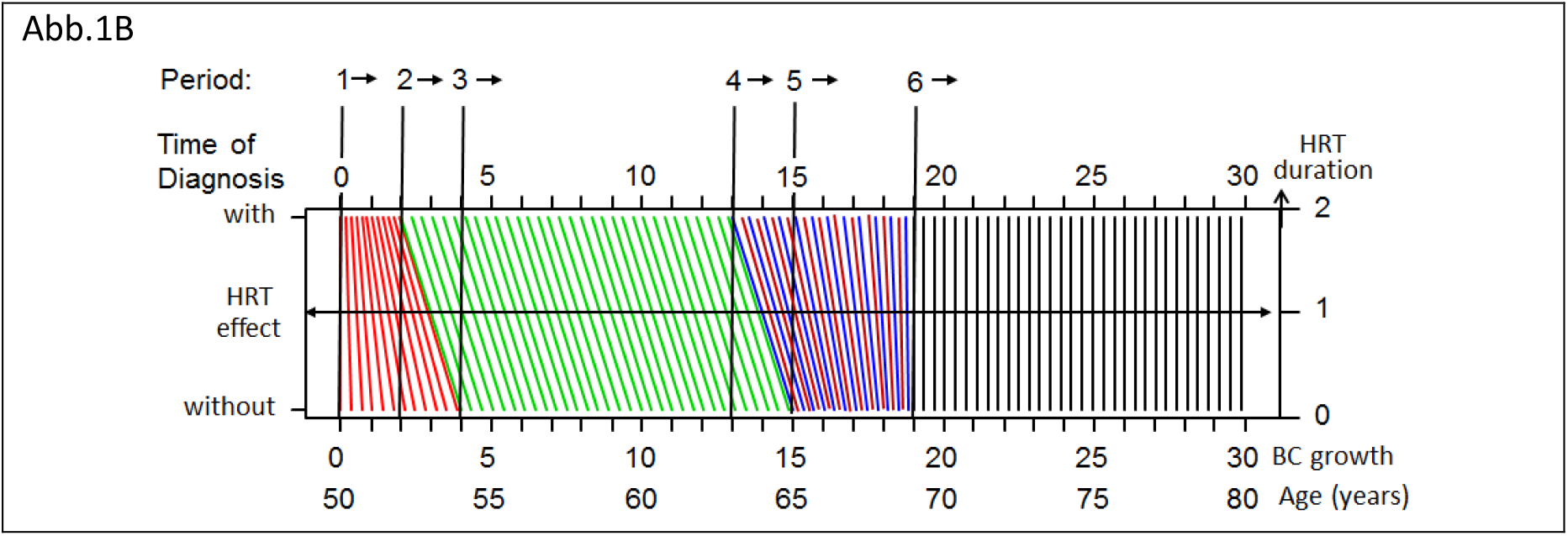
Tumor growth and the effects of hormone replacement therapy (HT) A: The x-axis represents age beginning at 50 years with a second x-axis representing the BC growth time relative to the beginning of the cohort. Double-sided arrows depict the time span of BC growth (black, mean 15 years) and the HT (4 years, red and gray area). On the left y-axis the number of tumor cells is shown (log-scale and example tumor diameters in mm), the right y-axis the age of the occult BCs. Their trajectories follow an exponential growth and are therefore shown as lines. Each trajectory depicts one of three respective growth spurts caused by HT (HTD of 4 years) with dashed lines representing the control group. The blue points at the top and bottom symbolize the origin of additional induced BCs and their shift in time to diagnosis after HT. (Periods: see Fig. 1B and legend). B: Diagnosis of BC with and without HT with a GD of 15 years, HTD 4 years, and a GF 2. The proliferation caused by HT initiates a transformation of the BC-free time span leading to 6 separate phases. The resulting concentration of BCs at the beginning leads to an apparently increased incidence. The 6 vertical lines distinguish time periods during follow-up: 1. HT beginning, 2. Turning point (HTD/GR), 3. End of HT, 4. Diagnosis of first BCs initiated or induced after start of HT and accelerated by HT, 5. Mean time of BC growth: Limit for all BCs prevalent at time=0. 6. End of HT effects (GD+HTD).

## Results

### Plausibility of model assumptions

For the model a GD of 15 years for ER+ BCs was assumed. Regarding chemoprevention, a rise in incidence in the following 15 years after the end of prevention was not found.^10-12^ This conservative presumption is presented in Figure 2A after a 2-year preventive therapy which successfully eradicated 50% of occult and BCs emerging on HTs. A GD of 15 years resulted in a duration of 166 days for the required 33 consecutive BC volume doubling of BCs of 20 mm.^13^ The GD was also estimated based on screening data.^14^ The calculated 25%/50%/75% percentile for the time required for a volume doubling in women between 60-69 years old and a tumor diameter of 15 mm was 65/143/308 days. Applying this to an increase in diameter from 2.5 mm (pT1a) to 28 mm (pT2) with 10.5 volume doublings resulted in 1.9/4.1/8.8 years. Based purely on this calculation and beginning with the first tumor cell, all BCs – not just ER+ – would grow to a diameter of 20 mm after 33 VDs in approximately 5.9/12.9/27.8 years.

**Fig 2:**
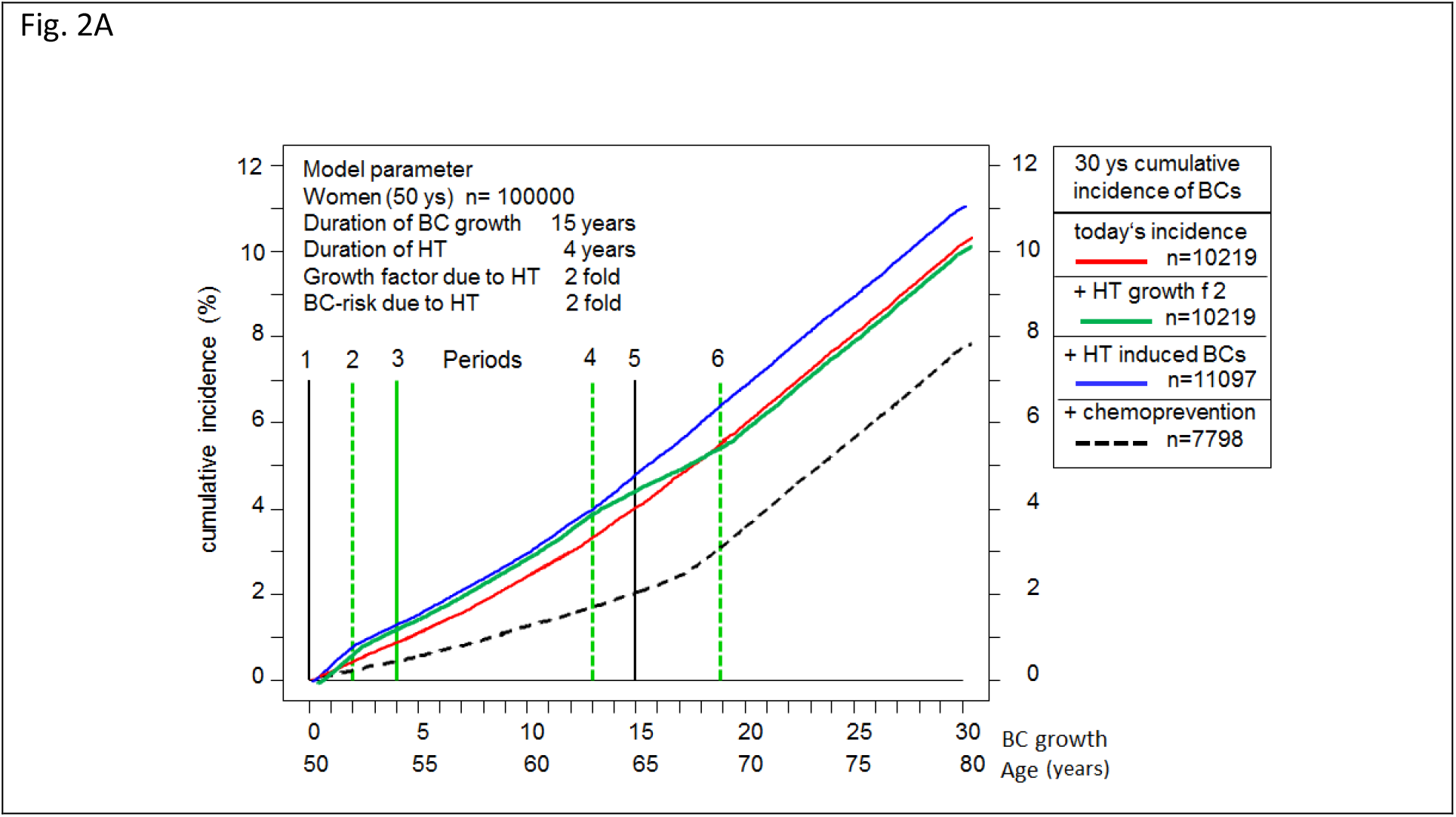
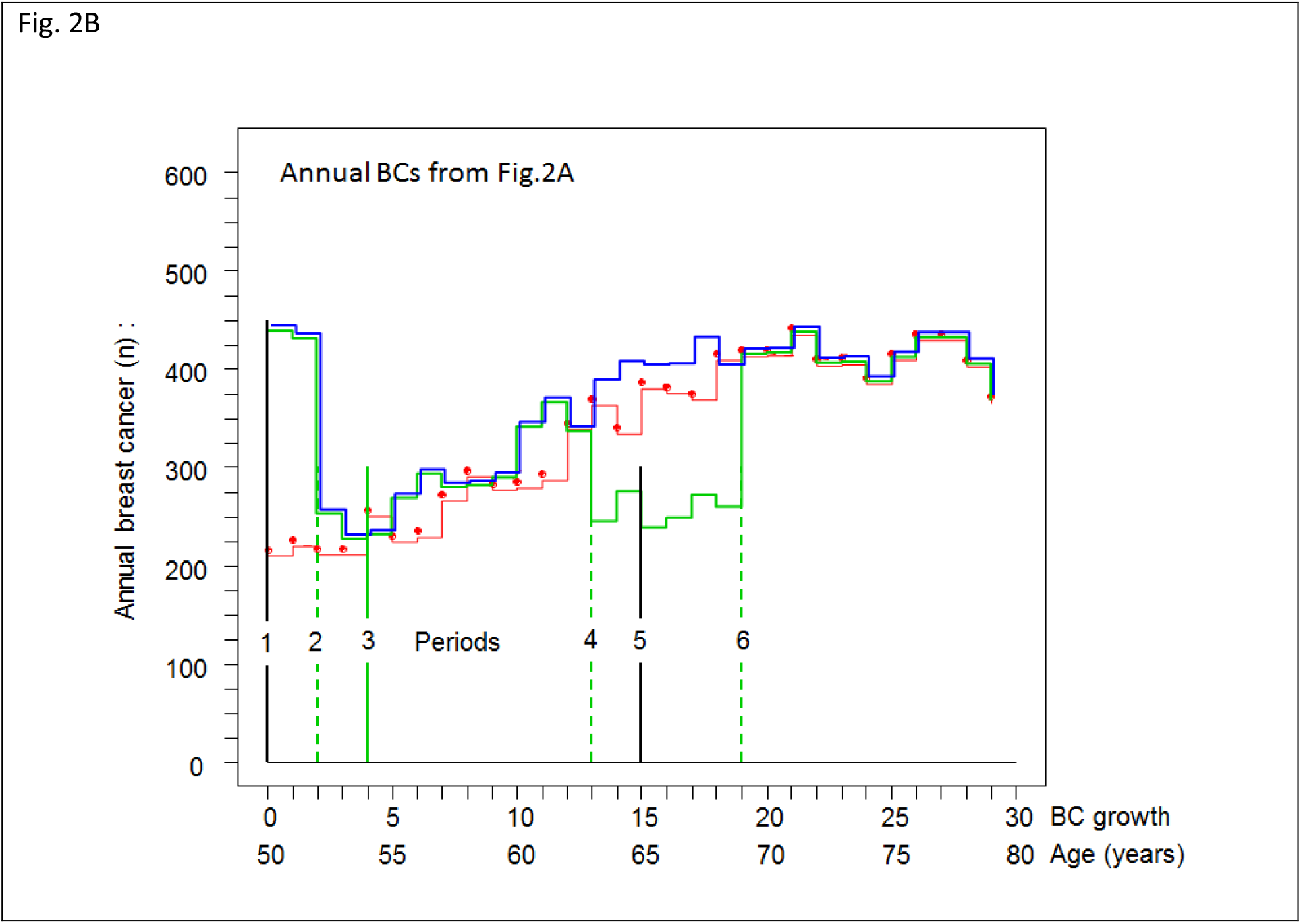
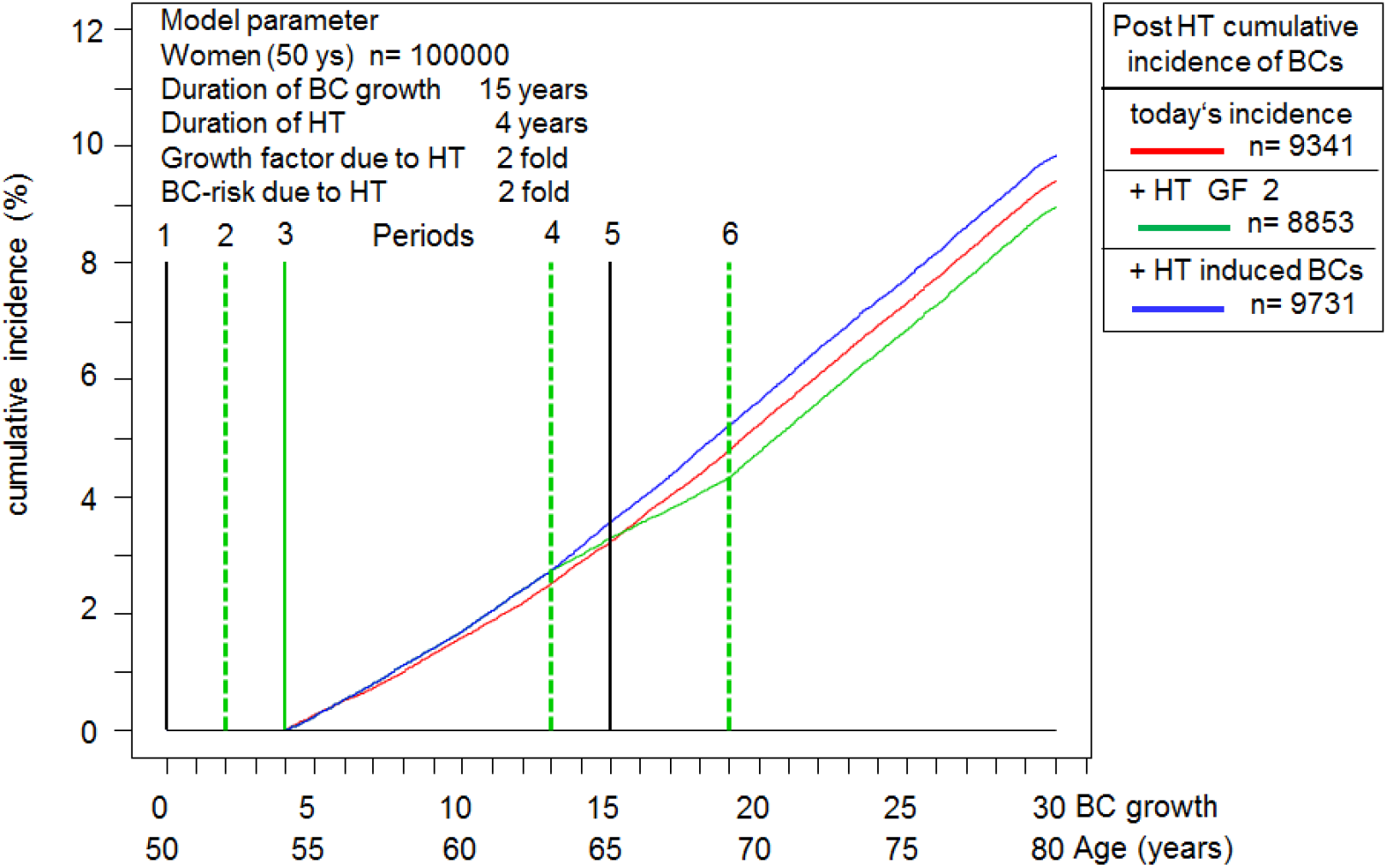
Simulation of the cumulative BC incidence for women starting from 50 years of age using the model parameters GD 15, HTD 4, GF 2, twofold BC risk. A: Cumulative BC incidence according to currently reported incidence (red), with accelerated growth after HT and the same number of BCs (green), and additional BC risk (blue). Were a cohort to receive a 50% effective chemoprevention for two years, the incidence would increase again two years after GD. Overlapping lines were manually shifted for better illustration. B: BC cases per year for each of the three cohorts of Fig. 2A without HT (red), with HT (green), and additional BC risk of HT (blue). The effect of GF 2 (green) with the concentration of cases in P1 (incidence doubling), the expedition of BCs, and the expansion in P4-5 are clearly evident for the same number of BCs. The HT induced BCs all occur during P4-5 (blue). Yearly fluctuations are caused by random generation. Overlapping lines were manually shifted to improve visualization. Fig.2C Cumulative incidence starting 4 years after HT end (post-HT cumulative incidence)

In addition a twofold BC risk was assumed for HT (Figure 2A), which by way of the Gail-Model^15^ estimated a higher prevalence of common risk factors. This model was used in prevention studies to recruit participants and confirmed its results ^11^. However, the GF in the model varied. In Figure 1 the factor of 2 is easily discernible and plausible following the above described doubling times. This GF was also identified from data on the 2.5-fold accelerated growth of ER- compared to ER+ BCs. In BC 20% of MET occur 10 years after the initial diagnosis, and together with the BC GD clarify for a time span of 30 years on the x-axis.^16^

### Simulation results

Results based on the parameters GD 15 years, HTD 4 years, and GF 2 are shown in Figure 1A. The trajectories illustrate a growth in diameter starting from BC initiation and the effect of HT. The model was based on the cumulative incidence of a cohort of 100 000 50 years old women (Figure 2A, red curve). Initially 216 BCs (SD 14) were randomly generated per year according to the age-specific SEER incidence, 30 years later the cases numbered 372 (SD 21) (Fig.2B), with an overall total of 10 219 (SD 105) BCs without considering competing risks. The slight curvature was a result of the increased incidence due to age, which is why only 40% of BCs occurred within the first 15 years. Chemoprevention applied over two years effective in 50% of cases could have prevented 2 431 or 24% BCs.

The application of HT stimulated the growth of occult BCs (Figure 2A, green curve). This lead to six marked follow-up periods P1-6 (Fig.1B) associated with HTD and GF. Three variations of this stimulated growth can be described, namely with 4 years of HTD with a GF of 2 there is a cumulation of BCs in the first two years, twice as many were diagnosed overall compared to the control group. Period P1 ends after two years (HTD/GF) at a turning point for the cumulative incidence. This point is defined by BCs that became apparent under HT 0 to a maximum of 2 years earlier following a linear course (Figure 1B P1 red with HT).

The second variation begins after 4 years when all BCs have been exposed throughout the HTD and become symptomatic two years earlier through this induced growth (Figure 1B green, P2-4 with HT). A gap subsequently appears in P4 (Figure 2B). In P4-6 (Figure 1B, brown) BCs were diagnosed also exhibiting accelerated growth that arise according the age-specific incidence for women aged 50 years or older under HT. The shift in this third variation spans from 0 to a maximum of two years similar to P1, however in the form of its mirror image. The consequences were as follows: The first BCs are maximally expedited and the duration of diagnosis is expanded thus lowering overall incidence. However, the cohort under HT (Figure 2A green) exhibits the same number, although faster growing, BCs as those without HT. Up until P6 the same number of BCs become evident and thereafter there is no longer a HT effect..

The concept that HT may induce de novo BCs was also simulated (Figure 1-2, blue). If HT caused BCs and these would be induced during HT with an accelerated growth rate, then these induced BCs would become apparent in the same phase P4-6 as the age-dependent BCs. A twofold BC-risk would double the number of normally HT induced BCs during an exposure to HT for women 54 years old or younger. A second turning point would be expected starting from P4 when the hazard rate would again increase due to BC risk (Figure 2C). It should be noted that HR at the 2nd turning point is likely to underestimate the true BC risk. If the growth of HT-induced BCs is also accelerated, then initiation occurs in P1-2 and diagnosis in the expanded period P4-6 (Fig.1B).

This type of non-proportional hazard rate can remain latent in an overall analysis. The hazard ratios (red vs. green) were identical up to the turning point due to the linear GF-transformation, only the confidence interval decreases due to the increasing number of patients. The HR is the quotient of the number of BCs that initially became evident with or without HT, therefore HR at this turning point is equivalent to the GF. After the turning point the HR fell to 1 in P6, meaning that even at the end of the HT the HR already underestimated the proliferative effect (HR 1.55 after 4 years in Figure 2A). If a BC risk is present (blue vs. red comparison) the HR may again increase from P4. Post interventional analysis (Figure 2C) can exhibit time lagged effects that may result in a protective effect in comparison to a cohort without HT.^17^

Due to the overlapping influence of BC risk and the proliferative effect small risks hardly become evident in studies with small sample sizes, a follow-up period spanning over 2 decades, and a lack of model assumptions. On the other hand it can be confirmed that the initial increase in incidence can be caused by the accelerated growth of previously existing BCs. Interpreting this as a BC-risk is therefore incorrect, it would be an incidence-increase imitation-bias.

### The women health initiative studies

In order to model the WHI-S the following data are used: Incidence, age-distribution, competing risks, and a sample cohort of 8 506 receiving HT with estrogen plus progestin for a total of 5.6 years. After 7.2 years the HR reached 1.24 (95%CI: 1.05 – 1.43).^8^ This HR can be achieved with a GF of 1.43 (95%CI: 1.18 – 1.78) with a turning point after 3.9 (5.6/1.43) years. The effect of the WHI-S can be explained with a boost in growth through HT (Figure 3). If after 13 years (HR=1.3, 95%CI 1.01-1.27) the HR continues to decrease, then there is no BC-risk with HT. Currently published is a HR of 1.29 (95%CI 1.14-1.47) based on 16.1 years of follow-up ^18^. This would correspond to a twofold BC risk after 19 years (HR 1.28, 95%CI 1.16-1.40).

**Fig. 3.**
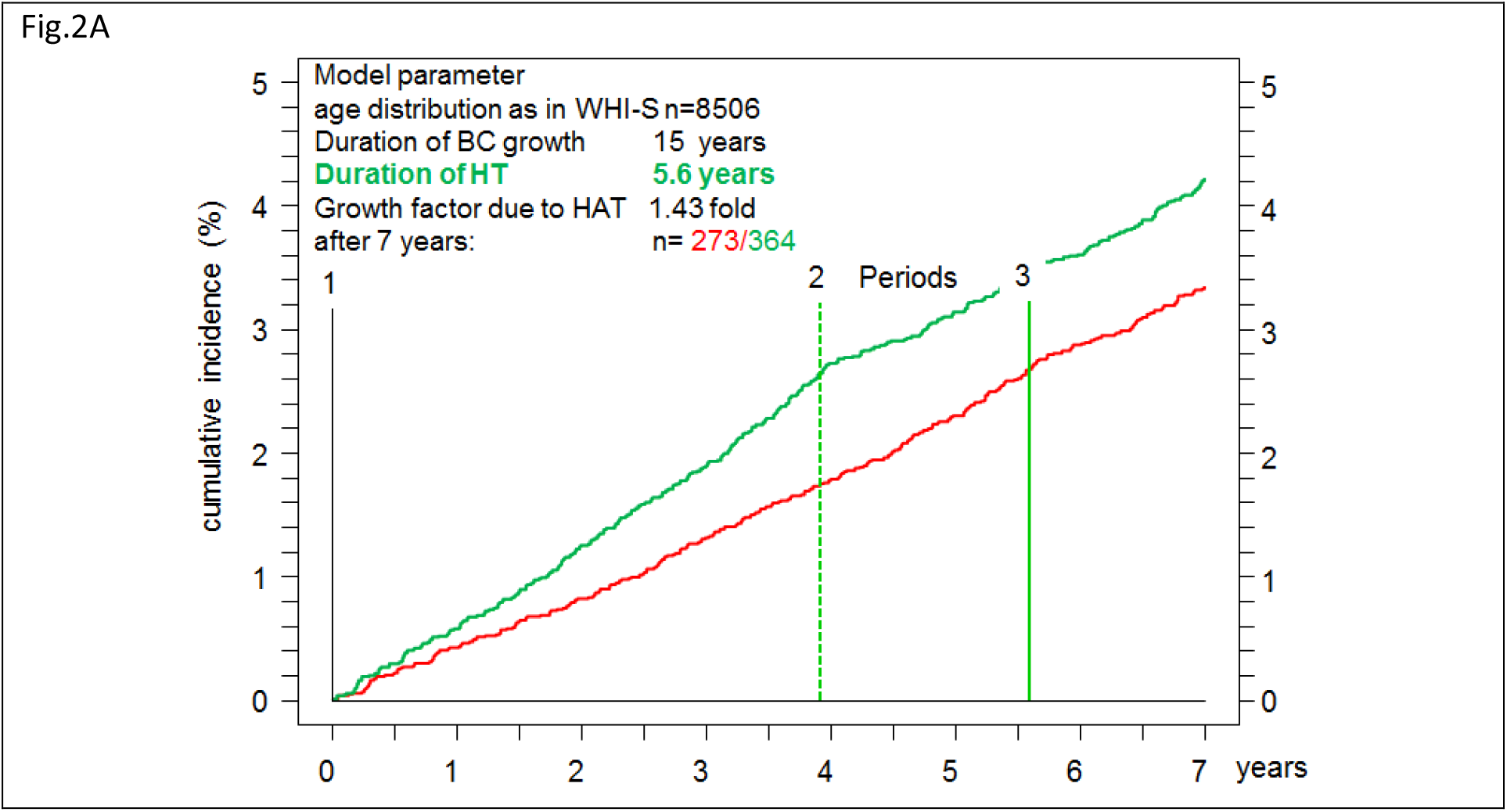
Simulations of the WHI-S using a cohort of n=8 506 according to their respective age-distribution (Mean age 63.7) and competing risks. HTD 5.6 years and GF 1.43 result in a HR of 1.26 after 7.2 years as reported in the WHI-S.

Even if the long-term results of the WHI-S were modeled with proliferation and BC risk, the discrepancy between the incidence curves should be noted. A negative mammography exam as an inclusion criterion could shift the first date of diagnosis of ER+ BCs by months, however a shift of 3 years as shown in the WHI-S is not plausible. This delayed opening of curves could not be reproduced in the WHI observational study which corresponds to Fig.3.^19^ A GF of 1.43 offered an explanation for the study results but did not directly explain the immediate drop in incidence observed in the United States after the publication of the study in July 2002. There was a 6.7% or 11.8% drop of BCs in all women or between the ages of 50-69 between 2002 and 2003 and the decrease was more evident in those with ER+ BCs.^5,20^

This simultaneous decline in the incidence of BCs and HT prescriptions can most likely be attributed to the publication of the associated BC risk. Simulations and the study data showed that HTs do lead to an apparent short-term increase in the incidence. This increase was just as brief and reversible as the change in breast density that results from HT discontinuation. Figure 1B illustrates the continued forward shift that resulted from a continued HT application. It should also be considered if no HT was started due to published BC risk, then no proliferative effect was induced accordingly. Taken together these results point to one reason for the observed drop in incidence, namely the decision of many women to abstain from or discontinue HT due to a perceived BC risk.

The simulation showed 8 506 women and 49/36 BCs in the first year with/without HT, in 100 000 women this added up to a total of 701/499 BCs. These results fluctuated because of the random generated data. Transferring these findings to the 203 500 newly diagnosed cases of BC in 2002 in the USA ^21^, out of which 44% were diagnosed between the age of 50-69, would explain the 11.8% drop in incidence equivalent to 10 570 BCs. A decline of this magnitude can be attributed to a GF of 1.43 and an average incidence of 360/10**5 in patients under HT if 6.8 million women discontinued or abstained completely from HT. A simulation of the WHI-S resulted in a difference of approximately 200 accelerated BCs out of 10**5 women after HT and 5.3 million women. However, the number of women under HT in total has decreased by 5 million between 2001 to 2003.^22^

### The meta-analysis

The updated meta-analysis summarized unique data from 58 clinical studies, included almost 150 000 BCs, and follow-up data of up to 20 years.^3^ Results herein for different HTD and post intervention phases were shown for the HT model with estrogen plus progestin. The remarkably high HRs for current users contradict these modeling results. A nearly implausible GF of 4 would still not be obtainable even after a HTD of 7 years and a resulting HR of nearly 2.

If the relative risk is determined using case-control studies the inclusion criteria must be observed in order to separate the two effects, growth acceleration and BC induction, according to Fig.2. Up to the turning point of the HT, the relative risk also estimates the GF. With an increasing time since HT in P2-P4, the HRs of past users kept becoming smaller compared to those of current users. If the sum of the HT duration and the time since the end of the HT reaches the mean GD of the BCs in P4-P6 for past users the BC risk can be analyzed. For several of these constellations, an increased BC risk of approximately 1.2 was shown after more than 10 years. An apparent protective effect due to proliferation (Fig. 2C red vs. green) cannot be seen, as in the WHI-S.

As with the WHI-S attention was also drawn to the increased BC mortality after 20 years for HT user. ^23^ The term proliferation was missing, so that an increased incidence did not differentiate between accelerated growth and BC risk.

## Discussion

The incidence and the associated BCs in the control group without HT are used as model parameters. The GD of ER+ BC varies greatly but 15 years appears to be a plausible assumption based on the exponential growth and volume doublings estimated from screening. The widening gap between incidence- and survival curves of chemoprevention and adjuvant therapy also highlight this observation. A GF of 1.43 explains the results of the WHI-S.^8,19^ The proliferation observed in HT exposed BCs characterised by a short-term increase in breast density and cellular activation is known from imaging during screening. This effect is reversible within a few weeks. Switching growth on and off through HT is the only plausible explanation for the sudden drop in BC incidence in 2002/2003.^5,24^ The change appears to be limited to the luminal A-like BC subtype and can be reversed by HT discontinuation.^25^ Similarly an increased level of Ki67 indicates an increased proliferation in benign breast tissue samples.^26^ This specific proliferative activity leads to a more frequent diagnosis of ER+ BCs under HT ^19^ compared to the ER- BCs 2.5 times faster growth rate.

While the HTs appear to accelerate short-term growth, the cellular pathways and characteristics of the induced BCs have so far not been adequately described. It is biologically plausible that BCs caused by HT are delayed.^27^ A risk relative to the age-specific incidence for women during HT was modeled. However, the assumption that the BC risk follows a linear increase and can be switched on and off similar to the proliferation activity is not plausible. Many risks do not follow a linear course, and at this time there is no evidence that HTs enable the rapid formation of the necessary driver mutations. This assumption would be in strong contrast to most other tumor types where carcinogenesis processes develop over years or even decades.^28,29^ By applying sensible assumptions the first BCs induced through HT would manifest at a later time, perhaps even 20 years after the start of HT.

In addition, data on the difference between BC-associated mutations promoted immediately through HT and those that occur normally as a result of serial mutations according to the known incidence remain to be reported. Are genetic modifiers of HT associated BCs available soon? Are there fewer passenger mutations due to short-term transformations in cancer cells? Do the 4% of occult BCs in 50 year old women present with a distinctive signature from the HT promoted growth? Do the BCs have an inherent memory for the exposition to HT? Is the growth boost dependent on tumor size? The molecular processes involved offer a unique research platform to study complex cancer pathways.^30^

Even epidemiological data currently do not offer sufficient evidence for HT-induced BC-risk.^31^ The chemoprevention results clarify that short-term eradication of prevalent BCs decreased the BC rate over more than 15 years.^10^ Similar results were seen for other risk factors such as alcohol consumption and smoking, which have an effect on cancer incidence after decades. However, up to this point it has not been proven that a withdrawal of HT can stop or even eradicate an already initiated cancerous growth. If the BC risk is efficiently reduced by millions of women who abstain from HT, then this should have a long-term effect on incidence. However, there was only a drop in incidence in 2002/2003 following the initial publication of the WHI-S and so far no further increase in the past 15 years.^5^ I.e. the status quo ante was already an artifact, the incidence has increased with the increasing use of HT, as the SEER data from 1995-2000 show. After the publication of the study results on the secondary prevention of coronary heart disease by HTs^32^ the BC incidence and the HT use have both continued to decline since 1999.^33,34^

The time with which BCs occur earlier increases continuously and reaches a maximum of HTD/GF years. The incidence in a population increases accordingly. If BCs grow 15 years, no induced BCs are expected in the first 10 years after the start of HT with a GF <1.5.

There is no valid evidence and it is not plausible^27^ that occult BCs shortly before becoming manifest should abandon their autonomous growth, regress, or only become evident at a later time through HT discontinuation.^20,35,36^ Even a slowly decreasing participation rate for screening examinations cannot explain this effect.^34,37^ The jump in incidence of about 10 000 BCs is a real effect and was achieved by about 5 million women not beginning or discontinuing an HT. With 360 BCs per 100,000 annually, this would achieve a short-term proliferation with a GF 1.59. A BC risk that only slowly increases with the duration of the therapy, as in meta-analysis, would lead to an implausibly high number of women who refrained from HTs or discontinued HTs. Since a GF of a specific HT should be uniform it would be desirable to prepare different HRs for the incidence curves when long-term follow-up is available.

Even if no evidence is available on a higher risk for recurrent cancer, then the entire process of cancerogenesis should be accelerated under HT. This includes the growth of BCs, tumor cell dissemination, and growth of prevalent metastases. Through this type of proliferation more cases of interval cancers, with a larger tumor size, increased number of node positive lymph nodes, and faster growing metastases occur as shown in the discussed studies.^8,23,38^ The 15-year survival of BCs with a diameter of 17 vs. 15 mm^8^ varies by 3%, a change from 11 to 15mm increases this difference to 5%.^39^ For this reason an annual instead of 2-3 yearly screening would have and will reduce proliferation risk in women with HT. A small increase in mortality is therefore plausible even without the additional BC risk.

Modeling the effect of a few parameters on the cumulative incidence and the post-interventional risk is a valuable approach. The interactions are complex, but in the sense of Occam’s razor, proliferation is a simple principle that can explain observations and stimulate the generation of hypotheses. For example the effect of a wash-out period of HTs on proliferation would be quite illogical. Rather, the sum of all HT-intervals should be correlated with the incidence. The increase in BC with prior HT use in the WHI-S has now been shown.^18^ Modeling studies can aid in continually specifying research objectives, prerequisites for future observations, and data analysis approaches. The previously discussed time-dependency and the P1-6 time-shifted effects are obstacles for any type of case-control studies. The immense value of prospective studies and real world data with follow-up data over decades becomes clear, since there is no valid data to point to specific factors which are correlated with frequency and timing of additional HT-induced BCs. This constitutes a major limitation: data is missing that allows models to be verified and improved.

The WHI-S showed that millions of women discontinued or completely abstained from HTs due a fear of cancer. By foregoing HTs many women suffering from postmenopausal symptoms have accepted a lower quality of life^2,40,41^. That should not be repeated. There are no valid estimates which of the 1 million BCs simply occurred earlier or which were induced with what timing. Balanced information also about possible induced and delayed BCs for women are necessary even after 50 years of HT.

## Conclusion

HTs appear to boost the growth of occult BCs, leading to an apparent rise in the overall incidence. A GF of 1.43 was estimated for the WHI-S which explains the decrease in incidence in 2002/2003. HT induced BCs would manifest simultaneous to the BCs originating under HT according to the age specific incidence. Although sound data on proliferation is available, type, timing, and especially the molecular characteristics of HT-induced rapidly growing BCs are currently not readily available. This work is mainly based on the analysis of cancer registry data and basic metastasis process. Modeling studies may aid in specifying research objectives and also provide direction during the search for effects in large datasets.

## Data Availability

The data used in this study is publicly available:
SEER agespecific incidence, lifetable (age specific normal mortality)
WHI-Study:n=8506, Logrank after 7.2 ys, age distribution

## Key clinical points

### Hormone Therapy for Postmenopausal Women

- The reported BC risk of HTs reduced their use after the publication of the WHI study in 2002 and caused an immediate 6.3% decrease in the incidence in the SEER data
- To date there is no reliable data on how large the BC risk is according to the type and duration of the HT
- Hormone receptor positive BCs grow slowly, on average maybe 15 or more years
- The increase in incidence with the onset of HTs can only be caused by a faster growth of occult hormone receptor positive BCs.
- Information is lacking on how quickly HTs induce the necessary driver mutations and whether these de novo BCs differ from normal BCs
- Induced BCs, even if mutations occur in a flash, are only likely to occur with a time delay of 10 or more years with growth factors < 1.5
- The 6.3% jump in incidence in a single year can only be caused by preventing faster growth
- The increased incidence in the SEER data up to this jump is correlated with the increasing HT use and caused by the assumed faster growth of occult BCs

#### Appendices

Disclosure The authors have declared no conflicts of interest.

## Acknowledgment

We thank Doris Mayr, Sylvia Heywang-Köbrunner, Christian Thaler, Michael Lauseker for valuable scientific and technical advice, Ingo Bauernfeind for the Project Group Breast Cancer for the opportunity to present and discuss preliminary results. Kathrin Halfter helped us with the manuscript and in particular, we would like to thank the staff of the Munich Cancer Registry for their meticulous and systematic recording and maintenance of the data.

## Abbreviations

BC: breast cancer
ER+/-: estrogen positive/negative
GD: Growth duration of BC
GF: Growth factor
HR: Hazard Ratio
HT: hormone replacement therapy
HTD: hormone replacement therapy duration
WHI-S: Women health initiative study

